# Diagnostic and prognostic value of alpha-synuclein seed amplification assay in Parkinson’s disease: a longitudinal cohort study

**DOI:** 10.1101/2024.12.03.24318422

**Authors:** Christina D Orrú, David P Vaughan, Nirosen Vijiaratnam, Raquel Real, Alejandro Martinez Carrasco, Riona Fumi, Marte Theilmann Jensen, Megan Hodgson, Christine Girges, Ana-Luisa Gil-Martinez, Eleanor J. Stafford, Lesley Wu, Bradley R Groveman, Andrew G Hughson, Olaf Ansorge, Annelies Quaegebeur, Kieren SJ Allinson, Thomas T Warner, Zane Jaunmuktane, Anjum Misbahuddin, P Nigel Leigh, Boyd CP Ghosh, Kailash P Bhatia, Alistair Church, Christopher Kobylecki, Michele TM Hu, James B Rowe, Thomas Foltynie, Huw R Morris, Byron Caughey, Edwin Jabbari

## Abstract

**Background:** Alpha-synuclein seed amplification assay (a-syn SAA) has been proposed to be a diagnostic biomarker for Parkinson’s disease (PD). Here, we have explored the diagnostic and prognostic value of cerebrospinal fluid (CSF) a-syn SAA status and seeding kinetics in PD.

**Methods:** Baseline CSF a-syn SAA data and longitudinal clinical data were collected and analysed between 1^st^ January 2010 and 1^st^ April 2022 for the Parkinson’s Progression Markers Initiative (PPMI) and UK parkinsonism cohorts respectively. We calculated the sensitivity and specificity of a-syn SAA in PD and controls, used linear regression to analyse a-syn SAA positive vs. negative group comparisons, and used time-to-event analyses to assess the ability of a-syn SAA seeding kinetic measures to predict clinical decline in PD.

**Findings:** We studied 1,402 participants: publicly available data from the PPMI cohort, n=1275 (PD, n=1,036; controls, n=239); newly generated data from the UK parkinsonism cohort, n=127 (PD, n=66; progressive supranuclear palsy (PSP), n=52; controls n=9). Over 2-5 years of follow-up, the sensitivity of a-syn SAA in PD was 87.7% and the specificity in controls was 91.9%. A-syn SAA was positive in 8/52 (15.4%) PSP samples with distinct ‘low and slow’ kinetics. A-syn SAA negative LRRK2-PD participants (n=57) had an older mean (SD) age at symptom onset (63.0 (7.6) vs. 55.4 (9.9) years) and higher mean (SD) baseline serum neurofilament light chain levels (20.4 (13.2) vs. 13.8 (8.6) pg/ml), p<0.05, vs. a-syn SAA positive LRRK2-PD participants (n=110). The baseline seeding kinetic measure, time to threshold, predicted cognitive decline in PD, defined as MoCA ≤21 (HR 2.51, 95% CI 1.50-4.20, p=0.001).

**Interpretation:** In PD, a-syn SAA may have value as a diagnostic and prognostic biomarker in clinical practice and as a stratification tool in clinical trials. Furthermore, we have highlighted the presence of pathological heterogeneity in LRRK2-PD.

**Funding:** Medical Research Council, PSP Association.

## Introduction

The differential diagnosis of parkinsonism includes Parkinson’s disease (PD), progressive supranuclear palsy (PSP), corticobasal syndrome (CBS), multiple system atrophy (MSA) and dementia with Lewy bodies (DLB). Each of these disorders is characterised pathologically by the prion-like spread of distinct structures of misfolded protein in specific brain regions leading to neurodegeneration and progressive neurological impairments.^1^ Misfolded neuronal alpha-synuclein (a-syn) in the form of Lewy body pathology is the hallmark of PD and DLB while MSA has a-syn pathology in the form of glial cytoplasmic inclusions.^2^ In contrast, PSP and corticobasal degeneration (CBD) are characterised by the presence of neuronal and astrocytic 4-repeat tau (4RT) pathology.^3^

In the absence of objective biomarkers, achieving an accurate diagnosis in patients presenting with symptoms suggestive of an underlying parkinsonian disorder is challenging, especially in the early stage of symptomatic disease when there is a high degree of clinical overlap between PD, PSP, MSA, CBD and DLB. This diagnostic challenge has been highlighted in previous cohort studies of patients who fulfil PD clinical diagnostic criteria where up to 10% go on to be re-diagnosed with alternative clinical diagnoses including PSP and MSA.^4^ Furthermore, there is significant variability in the rate of clinical disease progression in PD and this, combined with clinical diagnostic uncertainty, leads to a lack of homogenous groups for disease-modifying clinical trials.^5^

There is emerging evidence that a-syn seed amplification assays (SAA), which detect seeding-competent a-syn aggregates as a marker of Lewy body pathology, can reliably differentiate PD from healthy controls with high sensitivity and specificity, and may also be reliable in detecting pathology in the prodromal phase of PD and in differentiating PD from MSA.^6,7^ As such, there are growing calls to formalise a biological definition of neuronal a-syn disease for future research studies.^8,9^

Here, we have assessed the diagnostic and prognostic value of a-syn SAA status and seeding kinetics in PD by assaying the in-depth longitudinal UCL parkinsonism cohort and analysing publicly available data from the Parkinson’s Progression Markers Initiative (PPMI) cohort.

#### Research in context

##### Evidence before this study

We searched PubMed for articles on the diagnostic and prognostic value of CSF alpha-synuclein seed amplification assay (a-syn SAA) in Parkinson’s disease (PD) with no language restrictions from database inception up to November 1, 2024, using the following terms: “Parkinson’s disease AND CSF seed amplification assay”, “diagnosis OR prognosis”. Previous studies, including those involving the Parkinson’s Progression Markers Initiative cohort, have reported on the diagnostic value of a-syn SAA in PD based on cross-sectional analyses. However, the diagnostic value of a-syn SAA in the context of longitudinal stability of clinical diagnosis has not been explored previously. Similarly, previous analyses of the prognostic value of a-syn SAA seeding kinetics have been limited to small, single-centre, studies.

##### Added value of this study

To our knowledge, this is the largest study of the diagnostic and prognostic value of CSF a-syn SAA in PD. Although we highlight high sensitivity and specificity of a-syn SAA in PD and controls over a follow-up period of five years, our study is the first to demonstrate distinct ‘low and slow’ seeding kinetics in a-syn SAA positive CSF samples from progressive supranuclear palsy patients. We also highlight a distinct clinical and biomarker profile of a-syn SAA negative LRRK2-PD. Furthermore, our comprehensive time-to-event analyses have shown that the baseline a-syn SAA seeding kinetic measure, time to threshold (TTT), predicts cognitive decline in both sporadic and monogenic PD.

##### Implications of all the evidence available

A-syn SAA status may have value as a diagnostic biomarker in PD. However, the presence of a-syn SAA positivity with distinct ‘low and slow’ kinetics in a subset of PSP and PD samples reinforces the need for the development of 4-repeat tau SAA to be used in combination with a-syn SAA for greater diagnostic accuracy in clinical practice. Our results have also highlighted the presence of pathological heterogeneity in sporadic and LRRK2-PD, suggesting that a-syn SAA status could be used as clinical trial inclusion criteria. Furthermore, we have highlighted that TTT may be used as a prognostic biomarker and clinical trial stratification tool to recruit PD trial cohorts with homogenous disease progression trajectories.

## Methods

### Study design and participants

With regards to the PPMI cohort, data used in the preparation of this article were obtained on 2024-10-01 from the PPMI database (www.ppmi-info.org/access-dataspecimens/download-data), RRID:SCR_006431. For up-to-date information on the study, visit www.ppmi-info.org. Clinical and a-syn SAA data for sporadic PD, monogenic PD and control participants from the PPMI cohort were stratified according to whether CSF samples were assessed using the Amprion 24h or 150h a-syn SAA. Of note, previous reports of PPMI a-syn SAA data were based on samples analysed using the Amprion 150h a-syn SAA.^6^ Post-mortem data were not available on PPMI participants.

The UK parkinsonism cohort consisted of participants with PD, PSP and healthy controls who had provided written informed consent. PD participants fulfilling Queen Square Brain Bank clinical diagnostic criteria^10^ who were recruited to the Exenatide-PD3 clinical trial between January 1, 2020 to April 1, 2022 were included (ClinicalTrials.gov ID: NCT04232969).^11^ Only baseline (pre-drug/placebo) CSF samples were used as part of this study and post-mortem data were not available.

People with PSP fulfilling “possible” or “probable” clinical diagnostic criteria^12^ who were recruited to the PROSPECT-UK study’s natural history and longitudinal cohorts (Queen Square Research Ethics Committee 14/LO/1575) between September 1, 2015 to November 1, 2023 were included.^13^ Age-matched healthy controls who were recruited to the study were also included. Post-mortem evaluation was available on a subset of PSP participants.

### Procedures

Baseline CSF samples from participants in the PPMI cohorts were previously analysed using the Amprion 24h or 150h a-syn SAA as described.^6^

CSF sampling methods and application of the Rocky Mountain Laboratories (RML) a-syn SAA to baseline PD, PSP and age-matched healthy control CSF samples from the UK parkinsonism cohort are outlined in the **Supplementary Methods**.

In the UK parkinsonism cohort, matched baseline clinical data, i.e. obtained on the same day as the lumbar puncture, from PD and PSP patients consisted of sex, age at symptom onset, age and disease duration at baseline assessment, MDS-UPDRS-III score in the ‘Off’ state and MoCA score. Additionally, in PSP patients we noted the dominant PSP clinical subtype and the PSP rating scale score. PD participants had serial clinical assessment data available at 24-months post-baseline assessment and this was not stratified by drug/placebo status due to the Exenatide-PD3 trial not meeting its primary endpoint.^14^ Mortality and/or post-mortem data was not available on PD participants.

In the PPMI cohort, clinical data consisted of sex, age at symptom onset, age and disease duration at baseline assessment. Baseline, 2-year and 5-year scale scores for MDS-UPDRS-III in the ‘Off’ state, MoCA, SEADL and H&Y were recorded where available. Furthermore, change in clinical diagnosis and mortality status were noted on the date of censoring (November 1^st^ 2024).

Baseline neurofilament light chain (NFL) was measured in the plasma of PD patients in the UK parkinsonism cohort using the NULISA platform^15^ whereas baseline serum and CSF NFL was measured in a subset of PPMI cohort participants as described previously.^16^

In the UK parkinsonism cohort, ApoE status (rs7412 and rs429358), MAPT H1 status (rs1800547) and LRRK2-G2019S status (rs34637584) were defined using the KASP genotyping platform at the Laboratory of the Government Chemist (LGC Ltd), UK.

In the PPMI cohorts, genetic data from whole genome sequencing (WGS), whole exome sequencing (WES), GWAS, RNA-sequencing (RNA-seq), Sanger sequencing of select GBA variants, and mutation screening data (‘CLIA’) was obtained. This data was supplemented by ApoE and MAPT H1 status generated in-house at the PPMI, LRRK2, ADB, S4, and Bionet Biorepository at Indiana University. Variant data were extracted for the following genes using the same inclusion selection criteria as in the PD GENEration study (https://www.parkinson.org/understanding-parkinsons/causes/genetics/testing-counseling): LRRK2, GBA, SNCA, and PRKN. Variant data were reviewed to identify those that meet the current American College of Medical Genetics and Genomics criteria for pathogenicity, and data were compared across genetic platforms, to create a consensus variant resource for Parkinson’s disease researchers. The downloaded genetic data was the most up-to-date version released on 2023-12-20.

In samples assigned a positive a-syn SAA result, the following quantitative parameters of a-syn seeding kinetics were calculated from the ThioflavinT (ThT) fluorescence curves by calculating mean values of the replicates: time to threshold (TTT), the reaction time required to cross the fluorescence threshold; MaxThT, the maximum fluorescence during the reaction time; and AUC, the area under the ThT fluorescence curve integrated during the reaction time.

### Statistical analysis

Group comparisons were done using Fisher’s exact test for categorical variables and linear regression, adjusting for sex, age and disease duration at baseline, for continuous variables. For these comparisons, statistical significance was set at p < 0.05.

We used a time-to-event analysis to assess whether baseline a-syn SAA kinetic measures predict unfavourable outcome in a-syn SAA positive sporadic and monogenic PD participants. Building on previous studies,^4,17,18^ unfavourable outcome was defined as the development of either: a) cognitive decline (MoCA ≤21), postural instability (H&Y ≥3) or dependency (SEADL <80) during the 2 year post-baseline SAA testing follow up period in the UK parkinsonism cohort and the 5 year post-baseline SAA testing follow up period in the PPMI cohorts; b) death at any point from baseline SAA testing up until the date of censoring (October 1^st^ 2024). To harmonise all three assay cohorts for a well-powered analysis, in each individual cohort of a-syn SAA positive sporadic and monogenic PD participants we assessed each SAA kinetic measure separately and divided participants into two groups. In each cohort, the top quartile of MaxThT and AUC values were defined as “high seeding” groups and the bottom quartile of TTT values were defined as the “fast seeding” group due to having more aggressive seeding kinetics while the remaining participants were defined as “low seeding” and “slow seeding” groups. High/fast and low/slow seeding participants from each cohort were then combined for a Cox proportional hazards time-to-event analysis to assess whether each baseline a-syn SAA kinetic measure predicts unfavourable outcome, adjusting for sex, age and disease duration at baseline. For this analysis, a Bonferroni-adjusted p-value significance threshold was used.

All statistical analyses were performed in Stata, version 18 (StataCorp LLC) and GraphPad Prism 9.1.1 (San Diego, CA).

## Results

In total, publicly available data on 1,275 participants from the PPMI cohort were studied **(Table 1)**, with CSF samples previously analysed using the Amprion 24h or 150h a-syn SAA. Genetic status was available for 273/596 (45.8%) participants in the 24h Amprion a-syn SAA cohort and 679/679 (100%) participants in the 150h Amprion a-syn SAA cohort. PD participants without genetic data available were included in the sporadic PD group. The pathogenic variants detected in monogenic PD participants in the PPMI cohort are summarised in the **Supplementary Table 1**.

**Table 1:**
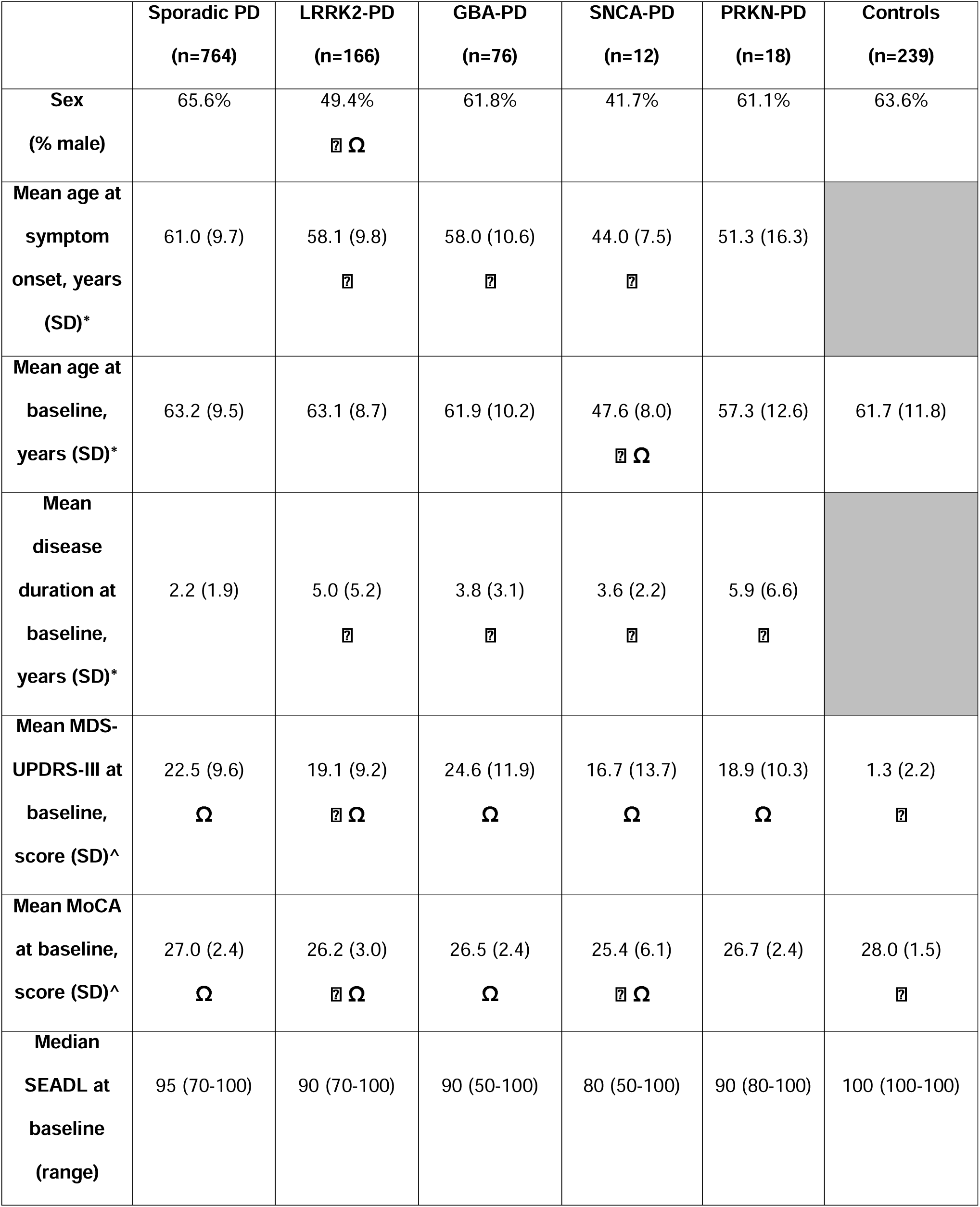
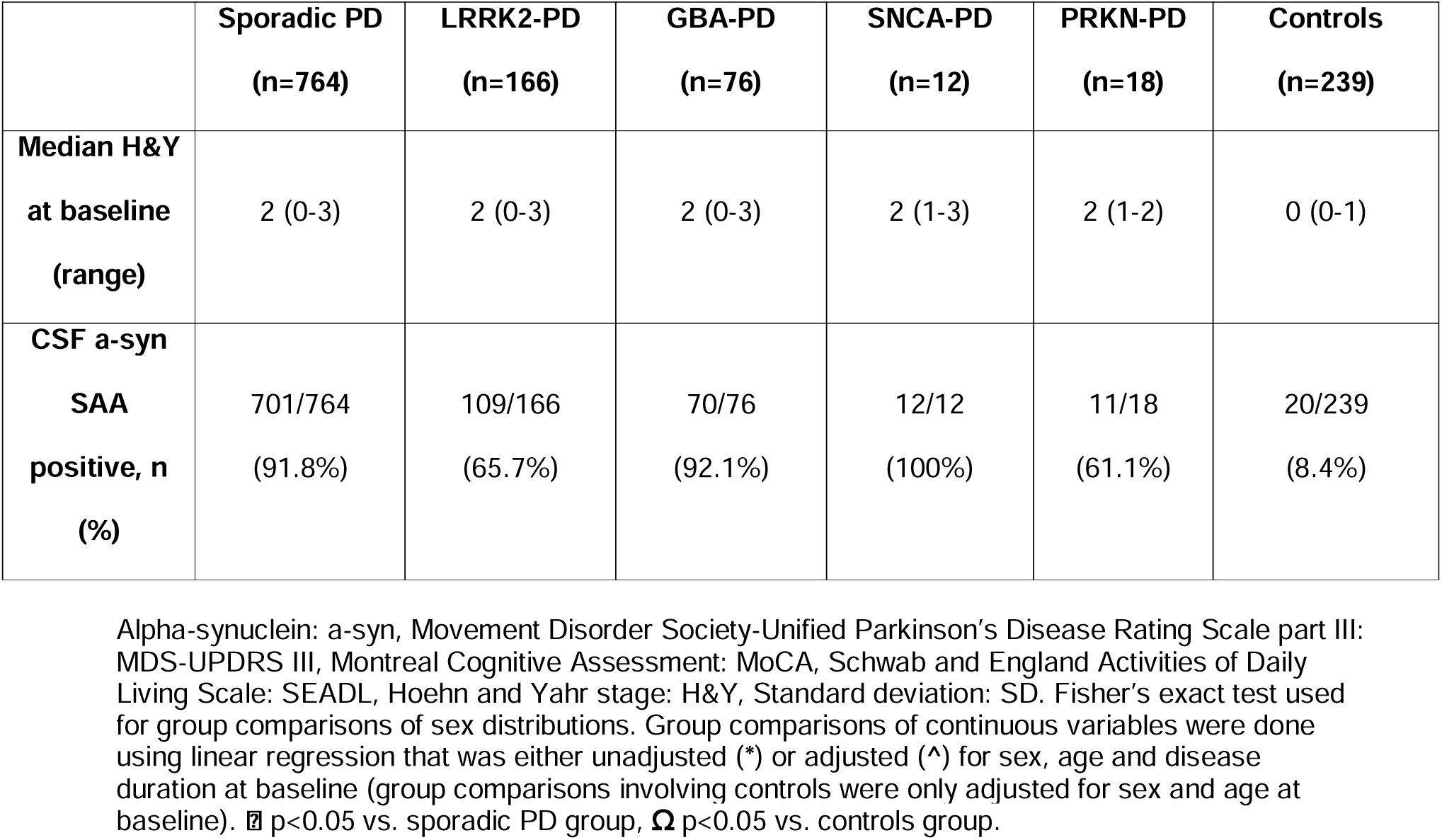
Baseline clinical profile and CSF a-syn SAA status of the PPMI cohort.

51 participants consisting of sporadic PD (n=28), LRRK2-PD (n=17), GBA-PD (n=4) and controls (n=2) had CSF samples analysed on both Amprion assays. Result concordance was high (47/51, 92.2%), with discordant results obtained in two sporadic PD and two LRRK2-PD samples. We found high rates of a-syn SAA positivity in sporadic PD (91.8%) and GBA-PD (92.1%) SNCA-PD (100%), whereas positivity rates were lower in LRRK2-PD (65.7%), PRKN-PD (61.1%) and controls (8.4%) **(Table 1)**.

66 participants with PD from the Exenatide-PD3 trial as well as 52 participants with PSP and 9 healthy age-matched controls from the PROSPECT-UK study were included in the UK parkinsonism cohort **(Table 2)**. Only one PD sample from the entire cohort tested positive for the LRRK2 G2019S variant.

**Table 2:** Clinical and CSF a-syn SAA profile of the UK parkinsonism cohort.

|  | PD (n=66) | PSP (n=52) | Healthy controls (n=9) |
| --- | --- | --- | --- |
| **Sex (% male)** | 78.8% | 59.6% $\Omega$ | 11.1% $\boxtimes$ |
| **Clinical subtype at baseline assessment (n)** |  | PSP-RS, n=31<br>PSP-P, n=10<br>PSP-PGF, n=3<br>PSP/CBS, n=3<br>PSP-F, n=3<br>PSP-SL, n=2 |  |
| **Mean age at symptom onset, years (SD) \*** | 52.6 (9.5) years | 64.4 (6.8) years<br>$\Omega$ | |
| **Mean age at baseline assessment, years (SD) \*** | 59.3 (8.6) years | 68.4 (6.7) years<br>$\Omega$ | 66.2 (3.9) years $\Omega$ |
| **Mean disease duration at baseline assessment, years (SD) \*** | 6.6 (3.6) years | 4.0 (2.1) years<br>$\Omega$ | |
| **Mean PSPRS at baseline assessment, score (SD) ^** |  | 34.3 (13.8) |  |
| **Mean MDS-UPDRS III at baseline assessment, score (SD) ^** | 33.5 (8.9) | 36.0 (14.0) |  |
| **Mean MoCA at baseline assessment, score (SD) ^** | 28.3 (1.5) | 22.2 (4.5) $\Omega$ | 28.8 (1.3) $\pounds$ |
| **CSF a-syn SAA positive, n (%)** | 63/66 (95.5%) | 8/52 (15.4%) | 0/9 (0%) |
| **Participants deceased at censoring (n), mean disease duration from symptom onset to death, years (SD)** |  | n=26<br>7.2 (3.0) years |  |
| **Primary pathological diagnosis in participants undergoing post-mortem (n)** |  | PSP, n=5<br>CBD, n=1 |  |

Corticobasal degeneration: CBD, Progressive Supranuclear Palsy: PSP, PSP-Richardson syndrome: PSP-RS, PSP-parkinsonism: PSP-P, PSP-progressive gait freezing: PSP-PGF, PSP/Corticobasal syndrome overlap: PSP/CBS, PSP-speech and language disorder: PSP-SL, PSP-frontal: PSP-F, number: n, standard deviation: SD, PSP rating scale: PSPRS, Movement Disorder Society-Unified Parkinson's Disease Rating Scale part III: MDS-UPDRS III, Montreal Cognitive Assessment: MoCA. Fisher's exact test used for group comparisons of sex distributions. Group comparisons of continuous variables were done using linear regression that was either unadjusted (\*) or adjusted (^) for sex, age and disease duration at baseline (group comparisons involving controls were only adjusted for sex and age at baseline). $\boxtimes$ = P<0.05 vs. PSP and PD groups, $\Omega$ = P<0.05 vs. PD group. $\pounds$ = P<0.05 vs. PSP group.

We applied the RML a-syn SAA in a blinded fashion to our CSF samples and found that 63/66 (95.5%) clinically diagnosed PD samples and 8/52 (15.4%) clinically diagnosed PSP samples were a-syn SAA positive **(Table 2)**. Of the 6 clinically diagnosed PSP participants who went on to have post-mortem evaluation: one had primary PSP pathology and Braak stage 1 Lewy body co-pathology and was CSF a-syn SAA positive; one had primary CBD pathology and Braak stage 3 Lewy body co-pathology and was CSF a-syn SAA negative; four had primary PSP pathology and no Lewy body co-pathology and were CSF a-syn SAA negative.

A plot of MaxThT vs. TTT values for all CSF a-syn SAA positive PD and PSP samples in the UCL parkinsonism cohort revealed a sub-group of positive samples with a distinct kinetic measure profile characterised by low MaxThT and prolonged time to threshold values, i.e. ‘low and slow’ kinetics, which we defined as samples that fell within both the bottom quartile of MaxThT values and the top quartile of TTT values in all a-syn SAA positive samples. Using these criteria, we found ‘low and slow’ kinetics in 8/63 (12.7%) positive PD and 6/8 (75%) positive PSP samples **(Figure 1)**. Relative to unequivocally a-syn SAA positive PD samples, a-syn SAA positive PD samples with ‘low and slow’ kinetics included individuals who were: a) older (>65 years) at symptom onset with a faster rate of motor progression; b) young (<40 years) at symptom onset with a similar rate of cognitive and motor progression **(Supplementary Table 2)**.

**Figure 1:**
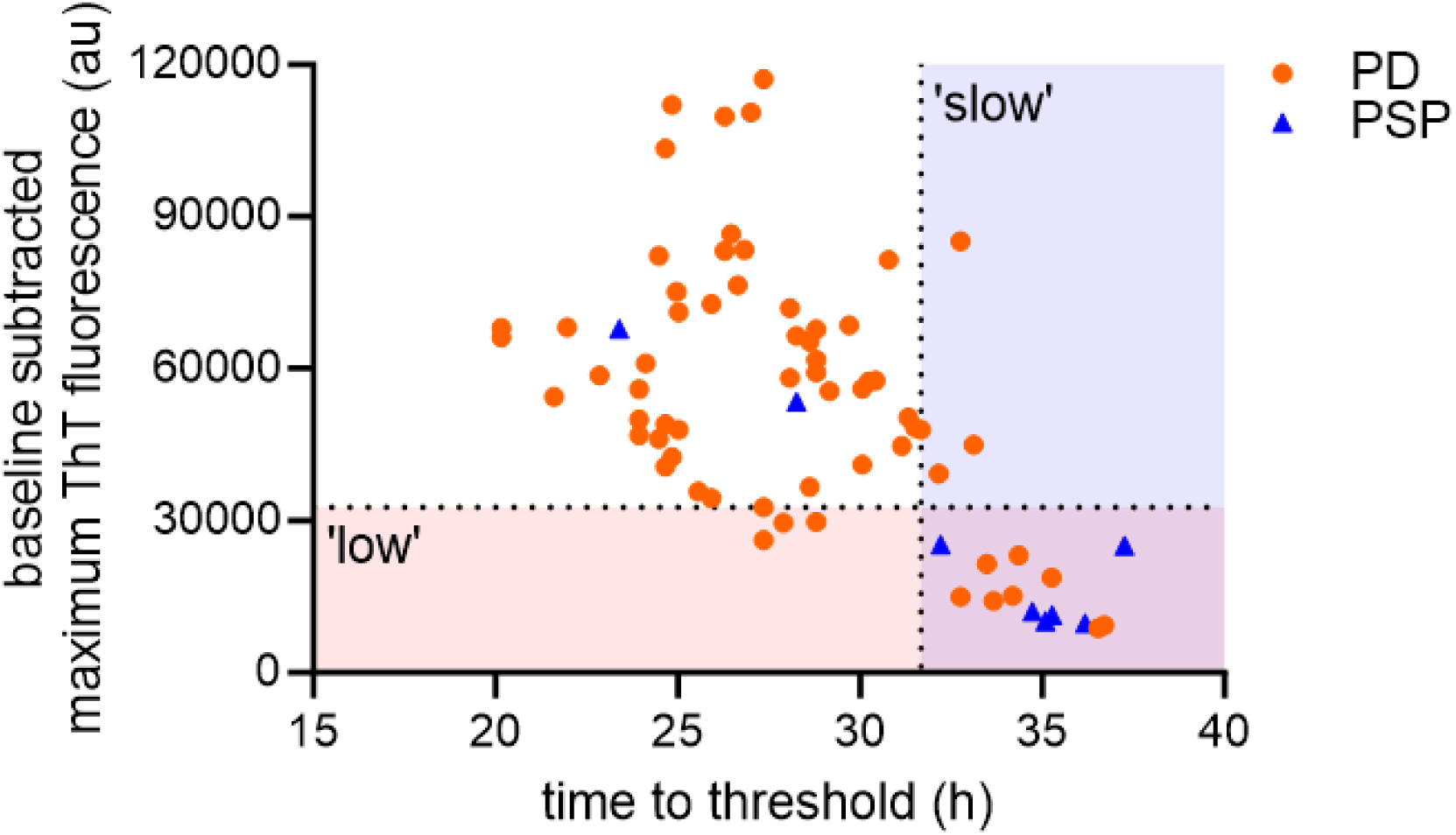
‘Low and slow’ kinetics of a-syn SAA positive CSF samples in the UK parkinsonism cohort. Cut-off points for the ‘low and slow’ group defined by the bottom quartile of both MaxThT values and the top quartile of TTT values in all a-syn SAA positive samples. Maximum ThT = Maximum thioflavin T fluorescence value; TTT = Time to threshold.

To calculate the overall sensitivity and specificity of a-syn SAA in clinically defined PD, we included sporadic and monogenic PD and control samples across all cohorts (total n=1,350). A negative result in a sporadic or monogenic PD sample was considered a false negative while a positive result in a control sample was considered a false positive. In the absence of pathological confirmation of diagnosis in PD participants, we factored in longitudinal change in clinical diagnosis since baseline SAA testing which was available over 2 years in the UK parkinsonism cohort and 5 years in the PPMI cohorts. Accordingly, only one PD participant from the UK parkinsonism cohort was clinically re-diagnosed as having DLB but their a-syn SAA positive result was considered a true positive due to the high likelihood of underlying Lewy body pathology. In the PPMI cohorts, two PD participants were clinically re-diagnosed as having MSA and so their a-syn SAA negative results were considered false negatives due to the high likelihood of underlying alpha-synuclein positive glial pathology.

Therefore, over 2-5 years of follow up, the sensitivity of the a-syn SAA in clinically defined PD was 87.7% and the specificity in controls was 91.9%.

We compared the clinical and biomarker profiles of a-syn SAA positive vs. a-syn SAA negative sub-groups within sporadic PD and LRRK2-PD groups across all cohorts. There was limited clinical follow up of a-syn SAA negative participants such that it was not possible to detect group differences in the rate of motor and cognitive progression.

In the LRRK2-PD analysis, the a-syn SAA negative group had an older age at onset and higher baseline serum NFL values in comparison with the corresponding a-syn SAA positive group **(Table 3)**. In the PPMI cohorts, we explored potential differences in a-syn SAA seeding kinetics in a-syn SAA positive monogenic PD vs. sporadic PD. We found that GBA-PD (in the 24h assay cohort) and SNCA-PD (in the 150h assay cohort) samples had a faster TTT relative to sporadic PD. In contrast, LRRK2-PD samples from the 150h assay cohort had a slower TTT relative to sporadic PD **(Supplementary Table 3)**.

**Table 3:** a-syn SAA positive vs. a-syn SAA negative analysis in sporadic PD and LRRK2-PD participants.

|  | Mean age<br>at<br>symptom<br>onset,<br>years (SD) | Mean 2-year<br>change in<br>MDS-UPDRS-III, n, score<br>(SD) | Mean 2-year<br>change in<br>MoCA, n,<br>score (SD) | Mean baseline<br>CSF NFL, n,<br>value pg/ml<br>(SD) | Mean baseline<br>serum NFL*, n,<br>value pg/ml (SD) |
| --- | --- | --- | --- | --- | --- |
| **Sporadic PD** |  |  |  |  |  |
| **a-syn**<br>**SAA**<br>**positive**<br>**(n=763)** | 60.1<br>(9.9) | n=487<br>+3.1<br>(9.7) | n=496<br>-0.4<br>(2.6) | n=211<br>119.6<br>(155.7) | n=332<br>14.7<br>(8.3) |
| **a-syn**<br>**SAA**<br>**negative**<br>**(n=66)** | 62.8<br>(10.0) | n=3<br>+1.3<br>(4.7) | n=3<br>+1.0<br>(1.0) | n=16<br>155.7<br>(75.0) | n=26<br>16.8<br>(8.3) |
| **LRRK2-PD** |  |  |  |  |  |
| **a-syn**<br>**SAA**<br>**positive**<br>**(n=110)** | 55.4<br>(9.9) | n=25<br>+6.7<br>(9.4) | n=24<br>+0.6<br>(2.1) | n=6<br>78.6<br>(21.0) | n=91<br>13.8<br>(8.6) |
| **a-syn**<br>**SAA**<br>**negative**<br>**(n=57)** | 63.0<br>(7.6)<br>‡ | NA | NA | n=2<br>114.8<br>(82.5) | n=47<br>20.4<br>(13.2)<br>‡ |

Alpha-synuclein: a-syn, Movement Disorder Society-Unified Parkinson's Disease Rating Scale part III: MDS-UPDRS III, Montreal Cognitive Assessment: MoCA, Schwab and England Activities of Daily Living Scale: SEADL, Hoehn and Yahr stage: H&Y, Neurofilament light chain: NFL, Not applicable: NA, Standard deviation: SD, Seed amplification assay: SAA. Group comparisons of continuous variables were done using linear regression that adjusted for sex, age and disease duration at baseline (group comparisons of age at symptom onset were only adjusted for sex). ‡ p<0.05 vs. corresponding a-syn positive group. \* NFL values restricted to those from PPMI participants.

We used a time-to-event analysis to assess whether baseline a-syn SAA kinetic measures predict unfavourable outcome as outlined in the Methods.

Unfavourable outcome was observed in 159/708 (22.5%) a-syn SAA positive sporadic and monogenic PD participants who had follow-up data available. Of note, none of the a-syn SAA positive sporadic PD participants from the UK parkinsonism cohort had reached unfavourable outcome at 2 years post-baseline a-syn SAA testing based on cognitive decline (MoCA ≤21) or death, but these samples were included in the analysis as censored data. A Cox proportional hazards model highlighted that only TTT reached nominal significance in predicting unfavourable outcome **(Figure 2a) (Supplementary Table 4)**. We then analysed each component of unfavourable outcome separately and found that TTT predicted cognitive decline over the 5-year follow-up period, reaching the Bonferroni significance p-value threshold, p<0.003 **(Figure 2b)**. We replicated this finding in sporadic PD participants only after excluding monogenic PD samples **(Figure 2c)**. We also evaluated the association of seeding kinetics with outcome using linear regression analyses in the PPMI a-syn SAA positive sporadic PD participants. This revealed that baseline TTT predicted 2-year change in MoCA score in both PPMI assay cohorts, while baseline AUC predicted 2-year change in MoCA score in only the Amprion 150h assay cohort **(Supplementary Table 5)**. An identical approach in the UK parkinsonism cohort revealed no significant associations.

**Figure 2:**
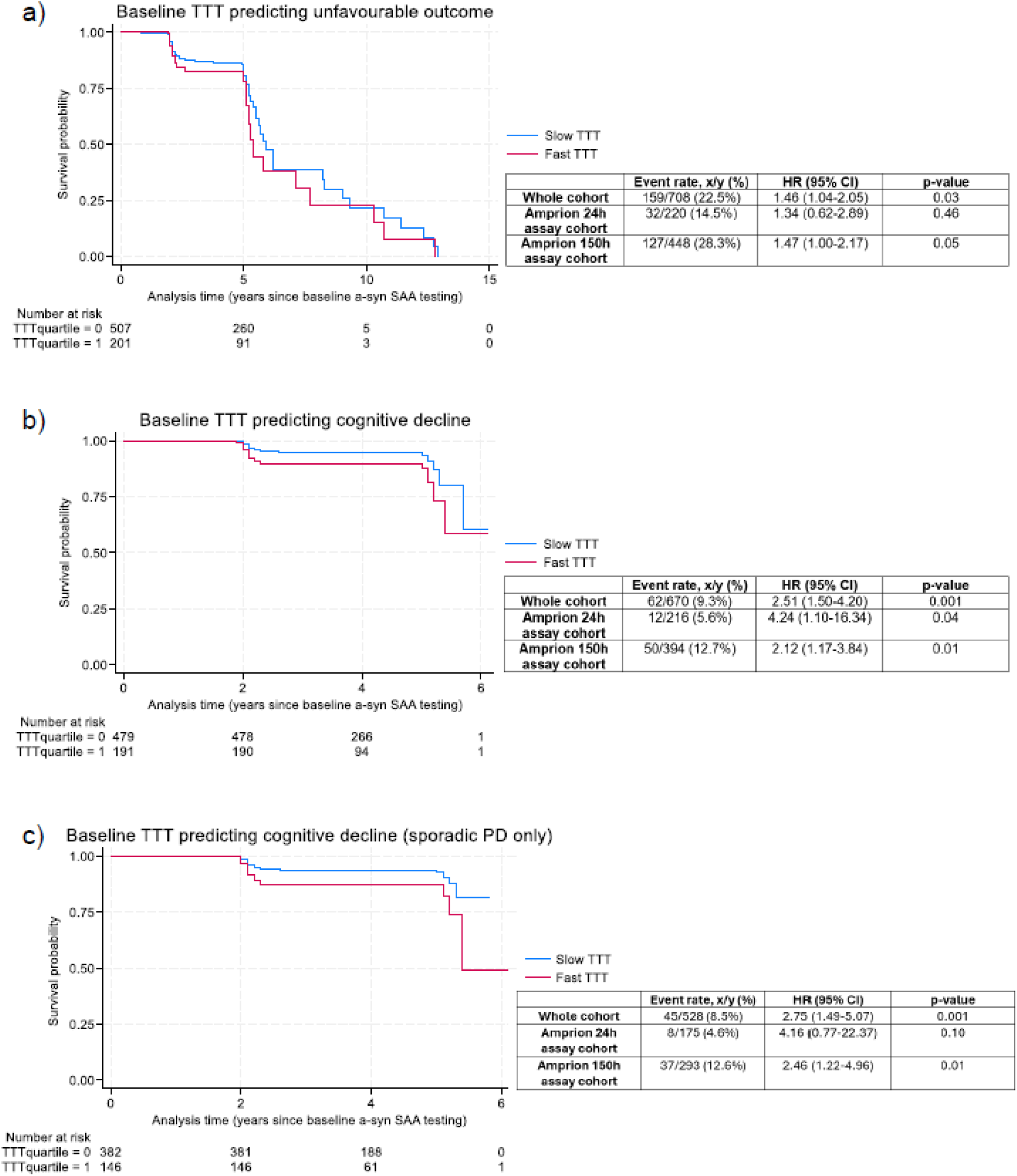
Time-to-event analyses stratified by baseline CSF a-syn SAA time to threshold. a) unfavourable outcome in sporadic and monogenic PD; b) cognitive decline in sporadic and monogenic PD; c) cognitive decline in sporadic PD only. Cox-proportional hazards time-to-event model used, adjusting for sex, age and disease duration at baseline. Associated summary statistics stratified by cohort included with each plot. Nominal significance p<0.05, Bonferroni significance p<0.003. a-syn SAA = alpha-synuclein seed amplification assay, TTT = Time to threshold, HR = hazard ratio, 95% CI = 95% confidence interval, Fast TTT = samples that are within the bottom quartile of TTT values, Slow TTT = samples that are within the top three quartiles of TTT values.

We found no significant associations between baseline a-syn SAA kinetic measures vs. serum and CSF NFL levels (p>0.05) in the PPMI cohort. In contrast, in the UK parkinsonism cohort we found significant associations between baseline plasma NFL vs. baseline MaxThT (r=0.37, p=0.005) and AUC (r=0.38, p=0.004) **(Supplementary Figure 1)**.

Across all three assay cohorts, we found no group differences (ApoE E4 vs. ApoE non-E4 and MAPT H1/H1 vs. MAPT non-H1/H1) in baseline a-syn SAA kinetic measures **(Supplementary Table 6)**.

## Discussion

In this study we have comprehensively assessed the diagnostic and prognostic value of a-syn SAA status and associated seeding kinetics in independent cohorts of PD participants, including the large-scale PPMI cohort of sporadic and monogenic forms of PD.

In line with previous a-syn SAA analyses of the PPMI cohort,^6,19,20^ our updated analysis with additional samples highlighted a high sensitivity and specificity of the assay in clinically-defined PD and controls, factoring in longitudinal stability of clinical diagnosis. Although different a-syn SAAs were used in our separate cohorts, we are reassured by previous work which has shown high concordance for results when PD CSF samples were applied to the Amprion 150h assay vs. the RML assay.^20^ Our current study has gone on to show high concordance for results comparing the Amprion 150h vs. 24h assay in CSF samples from sporadic and monogenic PD participants.

Our finding of a-syn SAA positivity in 15.4% of CSF samples from participants with clinically diagnosed PSP is in line with recent studies.^21,22^ We hypothesise that this represents Lewy body co-pathology as our a-syn SAA positivity rate is very similar to the rates of Lewy body co-pathology observed in post-mortem studies of PSP.^23,24^ This finding highlights that multiple proteinopathies may co-exist in individual patients and can be detectable during life. The presence of a-syn SAA positivity with ‘low and slow’ kinetics in both PSP and PD samples suggests that it is insufficient to rely on a-syn SAA positivity in isolation to support a clinical diagnosis of PD and/or provide biological evidence of neuronal a-syn disease as a stratification tool for clinical trial entry. Further refinement of the analysis of a-syn SAA kinetics may improve the performance of the assay in distinguishing PD from PSP. Furthermore, ongoing development of 4RT SAA^25,26^ is a major priority which will allow combined testing with a-syn SAA for more accurate stratification of patients with early-stage parkinsonism that is clinically indeterminate.

In the comparison of a-syn SAA positive vs. negative participants, our results suggest pathological heterogeneity within LRRK2-PD where the overall rate of a-syn SAA positivity in the PPMI cohort was 65.7%, in line with rates of Lewy body pathology detected in previous post-mortem studies of LRRK2-PD.^27^ With regards to potential alternative primary pathologies, there is an established link between LRRK2 and tau pathology, and most recently we have identified common variation at the LRRK2 locus as a genetic determinant of survival in PSP via a genome-wide association study of over a thousand PSP participants.^28^

Relative to sporadic PD, at baseline we found more aggressive seeding kinetics and a higher burden of cognitive impairment in GBA-PD and SNCA-PD whilst less aggressive seeding kinetics and a lower burden of motor impairment was observed in LRRK2-PD. This finding is in line with the differing rates of disease progression reported in these monogenic forms of PD.^29^

In the time-to-event analyses we found robust evidence of more aggressive a-syn SAA kinetics, as measured by fast TTT, predicting cognitive decline in PD over 2-5 years of follow up. This was apparent in monogenic PD, including GBA and SNCA mutations which are known genetic risk factors for dementia, but also in sporadic PD. Our finding is in line with a smaller separate longitudinal study of 199 sporadic PD participants which showed that baseline a-syn SAA kinetic measures predicted the development of cognitive impairment in PD (defined by MoCA score <25) with a mean study duration of six years.^30^ We hypothesise that higher a-syn SAA kinetic measures reflect higher seeding capacity which translates to a faster rate of cortical spread of Lewy body pathology and subsequent rate of cognitive disease progression. This suggests that a-syn seeding drives pathological and clinical disease progression and is therefore a justifiable disease-modifying target in PD. Furthermore, our results highlight that TTT may be used as a prognostic biomarker and trial stratification tool to recruit PD trial cohorts with homogenous disease progression trajectories.

Our study has some limitations that need to be considered. Post-mortem confirmation of neuropathology was only available in a subset of PSP participants so we cannot definitively comment on the sensitivity, specificity, positive predictive value and negative predictive value of a-syn SAA. Therefore, replication of our findings in larger clinical cohorts with post-mortem confirmation is desirable. The UK parkinsonism cohort was limited to genetic testing for only the LRRK2-G2019S variant, and comprehensive genetic status was established in only 45.8% of the PPMI Amprion 24h assay cohort, so it is likely that the sporadic PD groups in these two cohorts will contain a small number of unidentified monogenic PD cases which may influence our results.

Our results have highlighted high sensitivity of a-syn SAA in clinically-defined PD, although pathological heterogeneity may be especially relevant in LRRK2-PD. The presence of a-syn SAA positivity with distinct ‘low and slow’ kinetics in a subset of PSP and PD samples reinforces the need for the development of 4-repeat tau SAAs to be used in combination with a-syn SAA for greater diagnostic accuracy in clinical practice. Furthermore, our SAA kinetic data suggests that TTT may have a potential role as a prognostic biomarker in clinical practice and as a stratification tool in clinical trials for PD patients.

## Authors Contributions

Dr Jabbari (EJ) had full access to all the data and takes responsibility for the integrity of the data and the accuracy of data analysis.

DV, NV, RR, RF, MTJ, MH, AM, PNL, KPB, BCPG, AC, CK, MTMH, JBR, TF, HRM and EJ carried out study assessments for participants in the UK parkinsonism cohort; OA, AQ, KSJA, TTW and ZJ carried out post-mortem assessments on PSP participants from the PROSPECT-UK cohort; CDO, DV, NV, RR, AMC, RF, MTJ, MH, CG, ALGM, EJS, LW, BRG, AGH, CB, TF, HRM, BC and EJ contributed to the acquisition and analysis of data; EJ conceptualised and supervised the study; CDO and EJ drafted the manuscript; all authors critically reviewed the manuscript.

## Data sharing statement

De-identified data that support the findings and a data dictionary will be made available on reasonable request to the corresponding author.

## Acknowledgments

DV is supported by CBD Solutions. AQ is supported by Aligning Science Across Parkinson’s. CK is supported by the Multiple System Atrophy Trust and Parkinson’s UK. JBR is supported by the Medical Research Council (MC_UU_00030/14; MR/T033371/1), the PSP Association, Cambridge Centre for Parkinson-plus, the NIHR Clinical Research Network and the NIHR Cambridge Biomedical Research Centre (NIHR203312: the views expressed are those of the authors and not necessarily those of the NIHR or the Department of Health and Social Care). TF is supported by the National Institute of Health Research, Edmond J Safra Foundation, Michael J Fox Foundation, John Black Charitable Foundation, Cure Parkinson’s, Parkinson’s UK, Gatsby Foundation, Innovate UK, Janet Owens Research Fellowship, Rosetrees Trust, Van Andel Research Institute and Defeat MSA. HRM is supported by Parkinson’s UK, Cure Parkinson’s Trust, PSP Association, Medical Research Council and The Michael J Fox Foundation. EJ is supported by the PSP Association (PSPA2023/PROJECTGRANT001), CurePSP (681-2022/06) and the Medical Research Council (548211). The other authors have no funding declarations. This work was supported in part by the Division of Intramural Research of the NIAID. PPMI – a public-private partnership – is funded by the Michael J. Fox Foundation for Parkinson’s Research and funding partners, including 4D Pharma, Abbvie, AcureX, Allergan, Amathus Therapeutics, Aligning Science Across Parkinson’s, AskBio, Avid Radiopharmaceuticals, BIAL, BioArctic, Biogen, Biohaven, BioLegend, BlueRock Therapeutics, Bristol-Myers Squibb, Calico Labs, Capsida Biotherapeutics, Celgene, Cerevel Therapeutics, Coave Therapeutics, DaCapo Brainscience, Denali, Edmond J. Safra Foundation, Eli Lilly, Gain Therapeutics, GE HealthCare, Genentech, GSK, Golub Capital, Handl Therapeutics, Insitro, Jazz Pharmaceuticals, Johnson & Johnson Innovative Medicine, Lundbeck, Merck, Meso Scale Discovery, Mission Therapeutics, Neurocrine Biosciences, Neuron23, Neuropore, Pfizer, Piramal, Prevail Therapeutics, Roche, Sanofi, Servier, Sun Pharma Advanced Research Company, Takeda, Teva, UCB, Vanqua Bio, Verily, Voyager Therapeutics, the Weston Family Foundation and Yumanity Therapeutics.

