## Supplementary material for "Diagnostic and prognostic value of alpha-synuclein seed amplification assay in Parkinson’s disease: a longitudinal cohort study"

| **Section** | | **Page** |
| --- | --- | --- |
| **1)** | **Supplementary Methods: CSF sampling and a-syn SAA in the UK parkinsonism cohort** | **2** |
| **2)** | **Supplementary Table 1: Pathogenic variants detected in monogenic PD participants in the PPMI cohort** | **3** |
| **3)** | **Supplementary Table 2: Clinical profile of the a-syn SAA ‘low and slow’ PD samples in comparison with unequivocally a-syn SAA positive PD samples in the UK parkinsonism cohort** | **4** |
| **4)** | **Supplementary Table 3: Baseline kinetic profile of a-syn SAA positive samples in the PPMI cohort** | **6** |
| **5)** | **Supplementary Table 4: Baseline a-syn SAA kinetic measures predicting unfavourable outcome and individual components of unfavourable outcome (cognitive decline (MoCA ≤21), postural instability (H&Y ≥3) or dependency (SEADL <80) and death) across all cohorts** | **7** |
| **6)** | **Supplementary Table 5: Baseline a-syn SAA kinetic measures predicting 2-year and 5-year change in scale scores of motor, cognitive, and functional progression in a-syn SAA positive sporadic PD in the PPMI cohort** | **8** |
| **7)** | **Supplementary Figure 1: Scatter plots highlighting relationship between baseline MaxThT and AUC vs. plasma NFL in unequivocally a-syn SAA positive PD samples in the UK parkinsonism cohort** | **9** |
| **8)** | **Supplementary Table 6: Exploratory analysis of the impact of ApoE and MAPT status on baseline a-syn SAA kinetic measures and 2-year change in scale scores of motor, cognitive, and functional progression in a-syn SAA positive sporadic PD across all cohorts** | **10** |
| **9)** | **Supplementary References** | **12** |

**Supplementary Methods**

**CSF sampling and a-syn SAA in the UK parkinsonism cohort**

CSF samples from PROSPECT-UK PSP and control participants were centrifuged at 1750g at 4°C for 5 minutes within 15 minutes of lumbar puncture. The supernatant was then aliquoted into 500 microlitre aliquots before immediately storing at -80°C. In contrast, in Exenatide-PD3 PD participants the first 10 drops of obtained CSF were discarded and the remainder was collected and subsequently aliquoted into 1 millilitre aliquots before immediately storing at -80°C. Only aliquots with no previous freeze-thaw cycles were used for a-syn SAA testing in this study. Prior to testing, CSF samples were thawed and distributed into single-use ~65uL aliquots and re-frozen at -80°C. Single-use aliquots from each participant were then thawed for testing on the a-syn SAA assay.

The K23Q mutation of the αSyn sequence (Accession No. NM_000345.3) was engineered using Q5 Site-Directed Mutagenesis (NEB) using the primers CCACACCCTGTTGGGTTTTCTCAG and CAGAAGCAGCAGGAAAGAC, as previously described **(1)**, using a pET28 vector with an N-terminal His-tag (EMD Biosciences). The plasmid was transformed into BL21(DE3) Escherichia coli (EMD Biosciences). K23Q recombinant αSyn was purified as previously described with some modification. Briefly, following overnight auto-induction expression, a 1L Luria broth BL21(DE3) cell suspension was split into 4x250mL conical vessels and spun at 3273xg for 12min. Cell pellets were frozen at -80 °C for 20min and resuspended in 30ml of cold PBS per vessel. Next, each suspension was transferred into a 50ml falcon tube and probe sonicated in an ice bath (4 x 45sec with 15sec rests) at a power setting of 45% (Ultrasonic) to lyse the cells. Each tube was boiled for 20min and centrifuged at 9000xg for 60min. Then supernatants were combined and 50ml of Buffer A (20mM Tris pH7.4) were added. Next, the supernatant was filtered (0.22 μm vacuum filter) and loaded onto a 5mL Ni-NTA column (Cytiva 17525501). The middle section of the elution peak was collected, 20mL of Buffer A were added before moving onto a 5ml Q-HP (Cytiva 17115401) column using pre-chilled Buffers A and B2 (B2: 20mM Tris, 500mM Imidazole pH7.4). Following the collection of the middle peak fractions, the eluted material was filtered with 0.22 μm syringe filter and dialyze against pre-chilled PBS (3.5L overnight at 4 °C and another 3.5L PBS for 4 hrs the next day) using 3.5 kDa MWCO dialysis membrane (Thermo Scientific 68035). After another 0.22 μm syringe filtration, protein concentration was determined using a UV–VIS spectrophotometer and a theoretical extinction coefficient at 280 nm of 0.36 (mg/mL)^−1^ cm^−1^. Protein was dispensed in single use aliquots and stored at −80 °C.

UK parkinsonism cohort CSF samples were applied to an established a-syn SAA at the NIH Rocky Mountain Laboratories (RML).¹ Each well of a black 96-well plate with a clear bottom (Nalgene Nunc International) was preloaded with six glass beads (0.8 mm in diameter, OPS Diagnostics). Assays were set up in quadruplicate reactions per patient and seeded with 15 μL of CSF. Prior to addition of CSF, each SAA reaction mix was 85 μL of solution adjusted to give final reaction concentrations of 40 mM sodium phosphate buffer, 170 mM NaCl, 0.1 mg/mL K23Q recombinant αSyn (filtered through a 100 kD MWCO Pall filter, immediately prior to use), 10 μM thioflavin T (ThT) and 0.0015% sodium dodecyl sulfate (SDS). The plates were sealed (Nalgene Nunc International sealer) and incubated at 42°C in a BMG FLUOstar Omega plate reader. Plates were subjected to cycles of 1 min shaking (400 rpm double orbital) and 1 min rest for a minimum of 40 hours. ThT fluorescence measurements were taken every 45 min (450 +/− 10 nm excitation and 480 +/− 10 nm emission; bottom read). Each plate included a pool of non-diseased human CSF as a negative control and the same pool spiked with a 10^-5^ confirmed PD brain tissue dilution as a positive control (4 reactions each).

Fluorescence threshold for a positive reaction was calculated as 10% of the maximum value any well reached on each plate, which meant that the threshold was calculated individually for each plate, thus accounting for differences across experiments and plate readers. For a CSF sample to be considered positive, the fluorescence signal needed to exceed the threshold in at least 75% of replicate wells (e.g., ≥3 of 4) by the 40-hour reaction cut-off time. Samples that had no positive wells were assigned a negative result. Samples that were positive in 1 or 2 replicate wells underwent a repeat experiment and were only assigned a positive result if the fluorescence threshold was exceeded in ≥2 of 4 replicate wells in the repeat experiment.

**Supplementary Table 1:** Pathogenic variants detected in monogenic PD participants in the PPMI cohort

|  | **Amprion 24h assay cohort** | **Amprion 150h assay cohort** |
| --- | --- | --- |
| **LRRK2** | G2019S, n=38  R1441G, n=16  R1441C, n=1  I2020T, n=1  N1437H, n=1 | G2019S, n=109 |
| **GBA** | N409S, n=15  L483P, n=4  IVS2+1G>A, n=3  L29Afs*18, n=2  R502C, n=1  R159W, n=1  F216Y, n=1 | N409S, n=47  L483P, n=2 |
| **SNCA** | A53T, n=1 | A53T, n=11 |
| **PRKN** | Q25X / G430D, n=1  Q34Rfs*5 / Q34Rfs*5, n=1  p.Pro113fs, n=1  p.Gln34fs, n=1  Exon deletion, n=3 | R275W, n=4  Exon duplication, n=2  Exon deletion, n=2  P133_A134insP, n=1  P37L, n=1  R42P, n=1 |

Leucine-rich repeat kinase 2: LRRK2, Beta-glucocerebrosidase: GBA, Synuclein Alpha: SNCA, Parkin RBR E3 ubiquitin protein ligase: PRKN.

**Supplementary Table 2:** Clinical profile of the a-syn SAA ‘low and slow’ PD samples in comparison with unequivocally a-syn SAA positive PD samples in the UK parkinsonism cohort.

|  | **Age at symptom onset, years** | **Disease duration at baseline, years** | **MDS-UPDRS-III at baseline, score** | **2-year change in MDS-UPDRS-III, score** | **MoCA at baseline, score** | **2-year change in MoCA, score** |
| --- | --- | --- | --- | --- | --- | --- |
| **PD a-syn ‘low and slow’ 1** | 46 | 7.8 | 32 | +4 | 30 | 0 |
| **PD a-syn ‘low and slow’ 2** | 51 | 4.5 | 17 | +5 | 29 | 0 |
| **PD a-syn ‘low and slow’ 3** | 68 | 6.3 | 39 | +28 | 25 | -1 |
| **PD a-syn ‘low and slow’ 4** | 47 | 5.6 | 35 | NA | 26 | NA |
| **PD a-syn ‘low and slow’ 5** | 48 | 3.5 | 33 | +2 | 30 | 0 |
| **PD a-syn ‘low and slow’ 6** | 35 | 4.0 | 34 | +3 | 30 | 0 |
| **PD a-syn ‘low and slow’ 7** | 47 | 7.6 | 23 | NA | 29 | NA |
|  | **Age at symptom onset, years** | **Disease duration at baseline, years** | **MDS-UPDRS-III at baseline, score** | **2-year change in MDS-UPDRS-III, score** | **MoCA at baseline, score** | **2-year change in MoCA, score** |
| **PD a-syn ‘low and slow’ 8** | 49 | 6.8 | 31 | -3 | 30 | -1 |
| **PD a-syn positive group (n=55)*** | 53.4 (9.0) | 6.7 (3.9) | 34.1 (9.2) | +0.4 (7.2) | 28.3 (1.5) | -0.2 (1.4) |

Movement Disorder Society-Unified Parkinson’s Disease Rating Scale part III: MDS-UPDRS III, Montreal Cognitive Assessment: MoCA, * Values for the PD a-syn SAA positive group are expressed as mean (standard deviation).

**Supplementary Table 3:** Baseline kinetic profile of a-syn SAA positive samples in the PPMI cohort.

| **Amprion 24h a-syn SAA** | | | | | | |
| --- | --- | --- | --- | --- | --- | --- |
|  | **Sporadic PD (n=380)** | **LRRK2-PD (n=26)** | **GBA-PD (n=23)** | **SNCA-PD (n=1)** | **PRKN-PD (n=1)** | **Controls (n=14)** |
| **Mean Fmax, value (SD)** | 140909.8  (23274.0) | 144358.6  (22930.0) | 117429.9  (20979.0)  **♱ Ω** | 135485.3  (NA) | 135906.7  (NA) | 137810.3 (18237.1) |
| **Mean TTT, hours (SD)** | 10.1 (2.0) | 9.6 (1.9) | 9.2 (2.4)  **♱ Ω** | 7.7 (NA) | 7.9 (NA) | 11.2 (2.8) |
| **Mean AUC, value (SD)** | 5195143859.6  (979812982.0) | 5388230769.3  (692078125.0) | 4667639681.1  (1222145164.0)  **♱** | 5815333333.3  (NA) | 5565666667.0  (NA) | 4737968690.5  (1322221503.0) |
| **Amprion 150h a-syn SAA** | | | | | | |
|  | **Sporadic PD (n=321)** | **LRRK2-PD (n=83)** | **GBA-PD (n=47)** | **SNCA-PD (n=11)** | **PRKN-PD (n=10)** | **Controls (n=6)** |
| **Mean Fmax, value (SD)** | 85175.6  (25912.1) | 83432.5  (24209.0) | 89786.9  (23043.4) | 91649.8  (30839.5) | 70493.7  (25804.9) | 88443.7  (22577.7) |
| **Mean TTT, hours (SD)** | 65.6  (10.9)  **Ω** | 70.2  (14.4)  **♱** | 63.9  (11.1)  **Ω** | 54.3  (4.9)  **♱ Ω** | 66.3  (14.4) | 81.5  (20.3)  **♱** |
| **Mean AUC, value (SD)** | 26332577.3  (3719112.5)  **Ω** | 24397007.8  (4841043.6)  **♱** | 26739389.9  (3774203.8)  **Ω** | 30142730.3  (2398036.6)  **♱ Ω** | 25694147.3  (4443572.2) | 20898245.0  (6635823.3)  **♱** |

Alpha-synuclein: a-syn, Movement Disorder Society-Unified Parkinson’s Disease Rating Scale part III: MDS-UPDRS III, Montreal Cognitive Assessment: MoCA, Schwab and England Activities of Daily Living Scale: SEADL, Hoehn and Yahr stage: H&Y, Maximum thioflavin T fluorescence value: Fmax, Time to threshold: TTT, Area under the curve: AUC, Standard deviation: SD. Group comparisons of continuous variables were done using linear regression that was adjusted for sex, age and disease duration at baseline (group comparisons involving controls were only adjusted for sex and age at baseline). **♱** p<0.05 vs. sporadic PD group, **Ω** p<0.05 vs. controls group.

**Supplementary Table 4:** Baseline a-syn SAA kinetic measures predicting unfavourable outcome and individual components of unfavourable outcome (cognitive decline (MoCA ≤21), postural instability (H&Y ≥3) or dependency (SEADL <80) and death)) across all cohorts.

|  | **Fmax** | | **TTT** | | **AUC** | |
| --- | --- | --- | --- | --- | --- | --- |
| **Event**  **Event rate x/y (%)** | **HR**  **(95% CI)** | **p-value** | **HR**  **(95% CI)** | **p-value** | **HR**  **(95% CI)** | **p-value** |
| **Unfavourable outcome**  **159/708 (22.5%)** | 0.97  (0.65-1.45) | 0.88 | **1.46**  **(1.04-2.05)** | **0.03** | 1.40  (0.99-1.98) | 0.05 |
| **Cognitive decline**  **(MoCA ≤21)**  **62/670 (9.3%)** | 0.85  (0.46-1.58) | 0.61 | **2.51**  **(1.50-4.20)** | **0.001** | 1.55  (0.89-2.68) | 0.12 |
| **Postural instability**  **(H&Y ≥3)**  **45/640 (7.0%)** | 1.49  (0.76-2.94) | 0.25 | 1.09  (0.54-2.19) | 0.82 | 1.23  (0.61-2.47) | 0.57 |
| **Dependency**  **(SEADL <80)**  **82/638 (12.9%)** | 1.02  (0.60-1.73) | 0.94 | 1.54  (0.96-2.46) | 0.07 | 1.53  (0.95-2.46) | 0.08 |
| **Death**  **50/709 (7.1%)** | 1.07  (0.47-2.44) | 0.87 | 0.97  (0.50-1.88) | 0.92 | 1.03  (0.53-2.00) | 0.93 |

Hazard ratio: HR, 95% confidence interval: 95% CI, Montreal Cognitive Assessment: MoCA, Schwab and England Activities of Daily Living Scale: SEADL, Hoehn and Yahr stage: H&Y, Maximum thioflavin T fluorescence value: Fmax, Time to threshold: TTT, Area under the curve: AUC. Cox proportional hazards models that adjusted for sex, age and disease duration at baseline were used to predict whether “high/fast seeding” vs. “low/slow seeding” comparisons for each baseline a-syn SAA kinetic measure predicts the event. Nominal significance (yellow) p<0.05; Bonferroni significance (red) p<0.003.

**Supplementary Table 5:** Baseline a-syn SAA kinetic measures predicting 2-year and 5-year change in scale scores of motor, cognitive, and functional progression in a-syn SAA positive sporadic PD in the PPMI cohort.

|  | | **Amprion 24h a-syn SAA** | | | | |
| --- | --- | --- | --- | --- | --- | --- |
|  |  | **Mean baseline FMax** | | **Mean baseline TTT** | | **Mean baseline AUC** |
| **2-year change** | **MDS-UPDRS-III**  **(n = 149)** | Coeff = -94.64  p = 0.55 | | Coeff = -0.02  p = 0.11 | | Coeff = 2.20x10⁶  p = 0.73 |
|  | **MoCA**  **(n = 160)** | Coeff = -377.26  p = 0.52 | | **Coeff = 0.10**  **p = 0.03** | | Coeff = -4.84x10⁷  p = 0.05 |
|  | **H&Y stage**  **(n = 165)** | Coeff = -2118.96  p = 0.35 | | Coeff = -0.23  p = 0.22 | | Coeff = -1.80x10⁶  p = 0.99 |
|  | **SEADL**  **(n = 184)** | Coeff = 295.18  p = 0.05 | | Coeff = 0.00  p = 0.83 | | Coeff = 8.98x10⁶  p = 0.15 |
| **5-year change** | **MDS-UPDRS-III**  **(n = 36)** | Coeff = 35.21  p = 0.92 | | Coeff = 0.02  p = 0.41 | | Coeff = -7.63x10⁶  p = 0.56 |
|  | **MoCA**  **(n = 35)** | Coeff = -1890.64  p = 0.24 | | Coeff = 0.04  p = 0.70 | | Coeff = -7.46x10⁷  p = 0.24 |
|  | **H&Y stage**  **(n = 36)** | Coeff = -602.63  p = 0.92 | | Coeff = 0.53  p = 0.18 | | Coeff = -2.86x10⁸  p = 0.25 |
|  | **SEADL**  **(n = 37)** | Coeff = 285.55  p = 0.21 | | Coeff = 0.00  p = 0.92 | | Coeff = 6.67x10⁶  p = 0.50 |
|  | | **Amprion 150h a-syn SAA** | | | | |
|  |  | **Mean baseline FMax** | **Mean baseline TTT** | | **Mean baseline AUC** | |
| **2-year change** | **MDS-UPDRS-III**  **(n = 269)** | Coeff = 12.83  p = 0.94 | Coeff = 0.00  p = 0.97 | | Coeff = 9568.91  p = 0.66 | |
|  | **MoCA**  **(n = 270)** | Coeff = -684.26  p = 0.22 | **Coeff = 0.51**  **p = 0.02** | | **Coeff = -168189.80**  **p = 0.02** | |
|  | **H&Y stage**  **(n = 271)** | Coeff = -1639.06  p = 0.55 | Coeff = 0.42  p = 0.70 | | Coeff = 85980.50  p = 0.82 | |
|  | **SEADL**  **(n = 295)** | Coeff = 301.68  p = 0.12 | Coeff = -0.04  p = 0.63 | | Coeff = 14112.46  p = 0.60 | |
| **5-year change** | **MDS-UPDRS-III**  **(n = 224)** | Coeff = 13.43  p = 0.92 | Coeff = 0.05  p = 0.41 | | Coeff = -1930.86  p = 0.92 | |
|  | **MoCA**  **(n = 224)** | Coeff = -427.11  p = 0.39 | Coeff = 0.10  p = 0.63 | | Coeff = -41163.98  p = 0.56 | |
|  | **H&Y stage**  **(n = 225)** | Coeff = -2151.83  p = 0.42 | Coeff = 1.81  p = 0.11 | | Coeff = -469329.70  p = 0.22 | |
|  | **SEADL**  **(n = 263)** | Coeff = 59.88  p = 0.65 | Coeff = -0.05  p = 0.41 | | Coeff = 12750.52  p = 0.53 | |

Alpha-synuclein: a-syn, Linear regression coefficient: Coeff, Movement Disorder Society-Unified Parkinson’s Disease Rating Scale part III: MDS-UPDRS III, Montreal Cognitive Assessment: MoCA, Schwab and England Activities of Daily Living Scale: SEADL, Hoehn and Yahr stage: H&Y, Maximum thioflavin T fluorescence value: Fmax, Time to threshold: TTT, Area under the curve: AUC. Linear regression analyses that adjusted for sex, age and disease duration at baseline were applied to a-syn SAA kinetic measures and change in clinical scale scores. Nominal significance (yellow) p<0.05; Bonferroni significance p<0.0021.

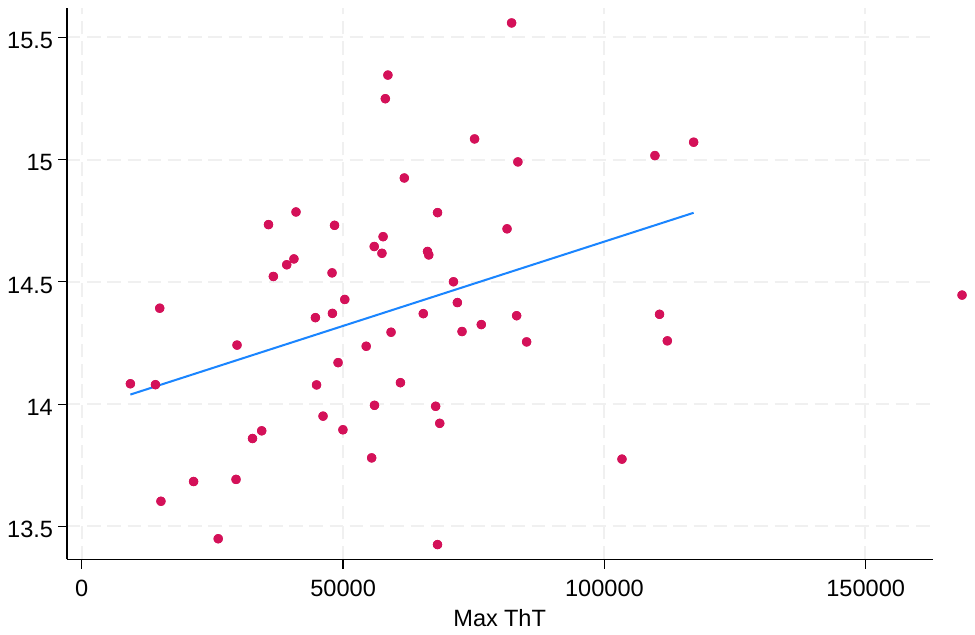
**Supplementary Figure 1:** Scatter plots highlighting relationship between baseline a-syn SAA kinetic measures vs. plasma NFL in a-syn SAA positive PD samples in the UK parkinsonism cohort.

r = 0.37

p-value = 0.005

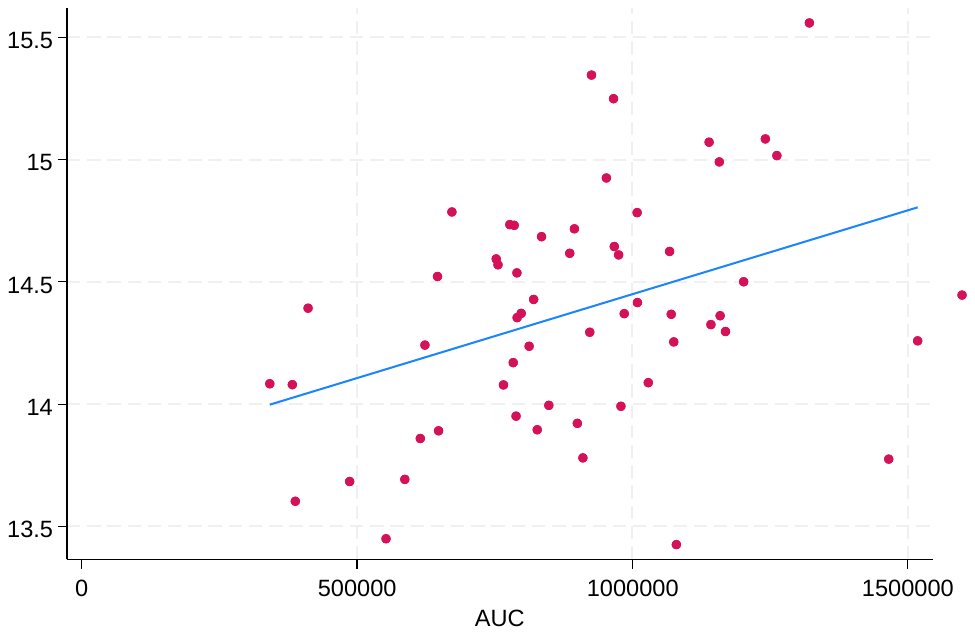

r = 0.38

p-value = 0.004

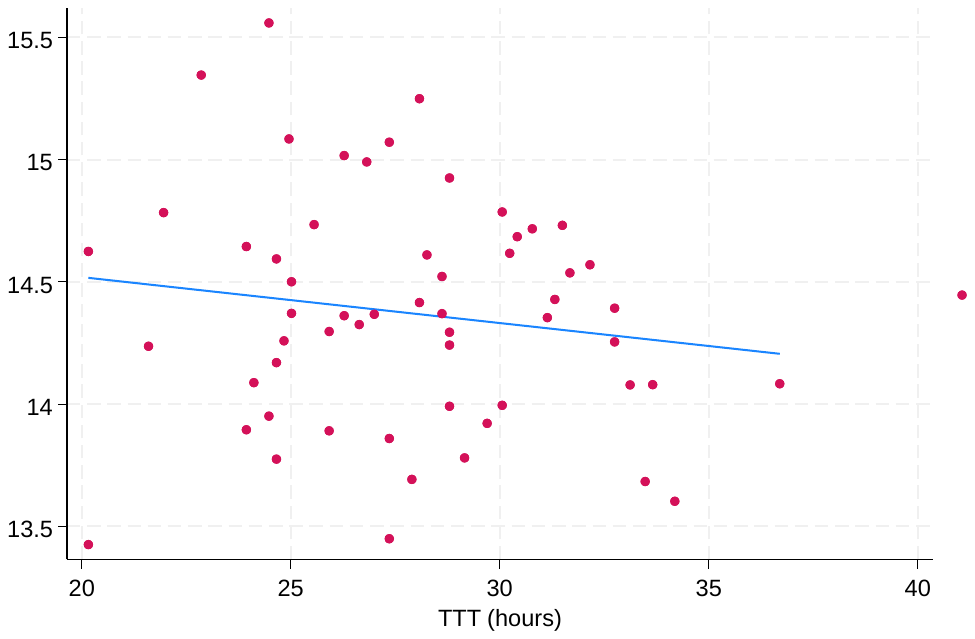

r = -0.15

p-value = 0.28

Alpha-synuclein: a-syn, neurofilament light chain: NFL, NULISA protein quantification unit: NPQ, Maximum thioflavin T fluorescence: MaxThT, Area under the curve: AUC. Linear trendline included in each plot.

**Supplementary Table 6:** Exploratory analysis of the impact of ApoE and MAPT status on baseline a-syn SAA kinetic measures and 2-year change in scale scores of motor, cognitive, and functional progression in a-syn SAA positive sporadic PD across all cohorts.

|  | | **Mean age at symptom onset, years (SD)** | **Mean 2-year change in MDS-UPDRS-III, n, score (SD)** | **Mean 2-year change in MoCA, n, score (SD)** | **Median 2-year change in SEADL, n, score (range)** | **Median 2-year change in H&Y, n, score (range)** | **Mean baseline FMax** | **Mean baseline TTT (hours)** | **Mean baseline AUC (SD)** |
| --- | --- | --- | --- | --- | --- | --- | --- | --- | --- |
| **UK parkinsonism cohort – RML a-syn SAA** | | | | | | | | | |
| **ApoE E4 carriers**  **(n=13)** | E3/E4, n=12  E4/E4, n=1 | 52.0  (8.9) | -4.1  (8.9) | -0.8  (1.3) |  |  | 53815.9  (16734.8) | 27.3  (3.3) | 852342.8  (173074.9) |
| **ApoE E4 non-carriers**  **(n=49)** | E2/E3, n=2  E3/E3, n=47 | 53.1  (9.4) | +2.4  (6.9) | -0.1  (1.4) |  |  | 56681.6  (27921.8) | 28.3  (4.0) | 878129.1  (290980.5) |
| **MAPT H1/H1**  **(n=36)** |  | 53.1  (9.0) | +0.4  (6.2) | 0.0  (1.4) |  |  | 53181.3  (24554.8) | 28.2  (3.5) | 850185.9  (256898.2) |
| **MAPT non-H1/H1**  **(n=26)** | H1/H2, n=22  H2/H2, n=4 | 52.5  (9.7) | +1.9  (9.8) | -0.5  (1.3) |  |  | 60095.5  (27598.9) | 27.8  (4.5) | 903926.4  (288232.5) |
| **PPMI cohort – Amprion 24h a-syn SAA** | | | | | | | | | |
| **ApoE E4 carriers**  **(n=35)** | E2/E4, n=1  E3/E4, n=30  E4/E4, n=4 | 61.3  (10.7) | +2.4  (9.6) | -0.3  (2.9) | -5  (-20 to +10) | 0  (0 to +2) | 130476.3  (19689.0) | 8.6  (0.9) | 5392466666.7  (872700651.0) |
| **ApoE E4 non-carriers**  **(n=90)** | E2/E2, n=2  E2/E3, n=13  E3/E3, n=75 | 59.4  (10.8) | +2.2  (8.9) | +0.2  (2.5) | 0  (-30 to +10) | 0  (-1 to +2) | 132835.9  (16386.4) | 8.8  (1.1) | 5310414814.8  (654181132.8) |
| **MAPT H1/H1**  **(n=32)** |  | 53.7  (13.3) | +4.6  (8.5) | -0.9  (3.0) | -7.5  (-25 to +5) | 0  (-1 to +2) | 137002.8  (18782.6) | 8.8  (1.0) | 5444447916.6  (755767352.5) |
| **MAPT non-H1/H1**  **(n=10)** | H1/H2, n=8  H2/H2, n=2 | 56.8  (9.8) | +2.2  (11.1) | -0.6  (2.0) | -5  (-20 to 0) | 0  (0 to +1) | 138144.2  (26809.1) | 9.3  (1.6) | 5319966666.6  (1021953822.8) |
| **PPMI cohort – Amprion 150h a-syn SAA** | | | | | | | | | |
| **ApoE E4 carriers**  **(n=89)** | E2/E4, n=9  E3/E4, n=72  E4/E4, n=8 | 57.8  (10.5)  **♱** | +5.2  (9.3)  **♱** | -0.7  (2.8) | 0  (-20 to +20) | 0  (-1 to +2) | 84813.9  (26679.3) | 66.0  (11.5) | 26201902.9  (3870770.5) |
| **ApoE E4 non-carriers**  **(n=246)** | E2/E2, n=3  E2/E3, n=36  E3/E3, n=207 | 60.6  (9.3) | +1.8  (10.2) | -0.5  (2.6) | -2.5  (-30 to +10) | 0  (-1 to +2) | 84524.6  (25460.5) | 65.5  (10.9) | 26269899.3  (3801347.1) |
| **MAPT H1/H1**  **(n=198)** |  | 59.4  (10.1) | +2.9  (9.7) | -0.7  (2.7) | 0  (-30 to +20) | 0  (-1 to +2) | 84371.3  (25351.8) | 65.3  (11.0) | 26370746.0  (3875020.5) |
| **MAPT non-H1/H1**  **(n=113)** | H1/H2, n=100  H2/H2, n=13 | 60.5  (9.4) | +3.2  (9.8) | -0.6  (2.5) | -5  (-30 to +10) | 0  (-1 to +2) | 83166.8  (24020.7) | 65.6  (11.2) | 26156324.7  (3762675.8) |

Alpha-synuclein: a-syn, Movement Disorder Society-Unified Parkinson’s Disease Rating Scale part III: MDS-UPDRS III, Montreal Cognitive Assessment: MoCA, Schwab and England Activities of Daily Living Scale: SEADL, Hoehn and Yahr stage: H&Y, Standard deviation: SD, Seed amplification assay: SAA, Maximum thioflavin T fluorescence value: Fmax, Time to threshold: TTT, Area under the curve: AUC. ApoE E4 vs. ApoE non-E4 and MAPT H1/H1 vs. MAPT non-H1/H1 group comparisons of continuous variables were done using linear regression that adjusted for sex, age and disease duration at baseline (group comparisons of age at symptom onset were only adjusted for sex). **♱** p<0.05 vs. corresponding genetic group.
